# INCIDENCE OF BENIGN PROSTATIC HYPERPLASIA IN TESTICULAR CANCER SURVIVORS IN THE VETERANS AFFAIRS HEALTH SYSTEM

**DOI:** 10.1101/2024.09.14.24313693

**Authors:** Kshitij Pandit, Kylie Morgan, Paul Riviere, Josue Cortez-Ryes, Margaret Meagher, Tyler Nelson, Dhruv Puri, Nuphat Yodkhunnatham, Frederick Millard, Rana R. Mckay, Brent Rose, Aditya Bagrodia

## Abstract

Testosterone and prostatic inflammation have been postulated to influence the development of benign prostatic hyperplasia (BPH). Our study aims to evaluate the incidence of BPH in Testicular Cancer (TCa) survivors, focusing on the impact of chemotherapy and post TCa diagnosed hypogonadism. We conducted a retrospective cohort analysis of US veterans diagnosed with TCa between 1990 to 2021, using the Veterans affairs database. BPH was defined using International Classification of Diseases (ICD) codes, Current Procedural Terminology (CPT) codes, or a 6-month prescription of medications. Associations with BPH were analysed, stratifying the cohort by receipt of chemotherapy and presence of hypogonadism. Multivariable cox regression models were used to determine statistical significance (p-value <0.05). Our cohort included 2038 TCa survivors with a median age at diagnosis of 41 years. On multivariable cox regression analysis, receipt of chemotherapy wasn’t significantly associated with incidence of BPH (p-value= 0.13). When stratified by diagnosis of hypogonadism prior to BPH, no significant associations were found on univariable (p=0.81) as well as multivariable (p=0.65) analyses. In the multivariable model, age at diagnosis was significantly associated with an increased incidence of BPH (Hazard ratio: 1.06, p<0.001). Our findings demonstrate that age is a significant factor associated with development of BPH in this population, while suggesting that chemotherapy for TCa and hypogonadism might not substantially alter the development of BPH.

**Patient summary:** In this study, we looked at whether testicular cancer survivors are more likely to develop an enlarged prostate (Benign Prostatic Hyperplasia), especially if they received chemotherapy or had low testosterone afterward. We found that age, rather than cancer treatment or testosterone levels, was the main factor linked to developing an enlarged prostate.

## Main report

Testicular cancer (TCa) is the most common malignancy among young men, and with improved survival rates, attention has shifted toward the long-term health impacts of cancer treatment^1^. Benign prostatic hyperplasia (BPH) is a common condition affecting men, characterized by lower urinary tract symptoms that significantly affect the quality of life^2^. While testosterone and prostatic inflammation are established factors in BPH development^3,4^, the potential influence of chemotherapy and hypogonadism on BPH in TCa survivors remains unclear.

Our study aims to evaluate the incidence of BPH in a cohort of TCa survivors within the Veterans Affairs Health System, with a specific focus on the potential association of chemotherapy and post-TCa hypogonadism with BPH. By understanding these associations, we seek to contribute to the development of targeted survivorship care strategies that address the unique health needs of TCa survivors.

### Study Design and Population

We conducted a retrospective cohort study using data from the Veterans Affairs (VA) Health System. The study population included U.S. veterans diagnosed with TCa between 1990 and 2021. Eligible patients were identified from the VA Cancer Registry, and those with incomplete data or incorrect histology were excluded. The final cohort consisted of 2,038 TCa survivors.

### Definition of Outcomes

The primary outcome was the incidence of BPH, defined using International Classification of Diseases (ICD) codes, Current Procedural Terminology (CPT) codes, or a six-month prescription of BPH-related medications. Procedures indicative of BPH included transurethral resection of the prostate (TURP), laser enucleation, simple prostatectomy, and Rezūm. Medications included were α-blockers, 5α-reductase inhibitors, anticholinergic agents, and β-3 agonists^2^ (included in supplementary material).

### Exposure Variables

The main exposure variables were chemotherapy and hypogonadism following TCa diagnosis. Hypogonadism was identified through ICD codes or testosterone prescriptions lasting six months or longer (supplementary material). We tabulated whether patients received chemotherapy as a part of their treatment course through VA cancer registry data.

### Statistical Analysis

Associations between chemotherapy, hypogonadism, and BPH incidence were analysed using multivariable Cox proportional hazards regression models. Covariates in the models included age at TCa diagnosis, race, smoking status, and clinical stage. Patients were stratified by chemotherapy receipt and hypogonadism diagnosis to assess their associations with BPH development. Statistical significance was defined as a p-value <0.05.

The study included 2,038 testicular cancer (TCa) survivors, with a median age of 42 years at diagnosis. Of these, 685 (33.6%) received chemotherapy, and 365 (17.5%) were diagnosed with hypogonadism after their TCa diagnosis but before a BPH diagnosis (Table 1). The incidence rate of BPH in TCa survivors was 21% at 10 years and 31% at 15 years. On multivariable Cox regression analysis, chemotherapy was not significantly associated with BPH incidence (p=0.13). Similarly, hypogonadism was not significantly associated with incidence of BPH in both univariable (p=0.9) and multivariable (p=0.63) analyses. Age at TCa diagnosis was significantly associated with BPH incidence (HR: 1.06, 95% CI: 1.05-1.07, p<0.001) (Table 2).

**Table 1:**
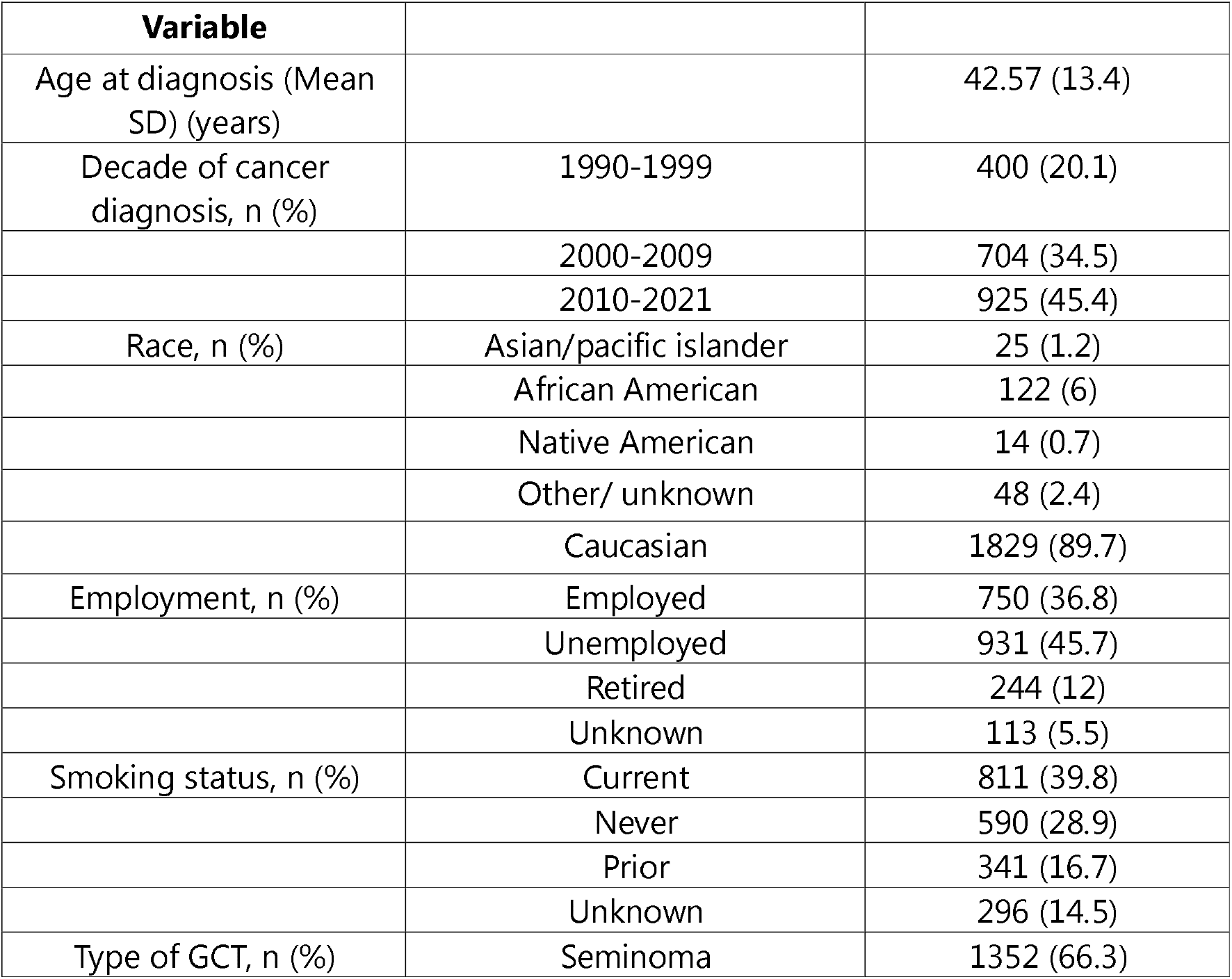
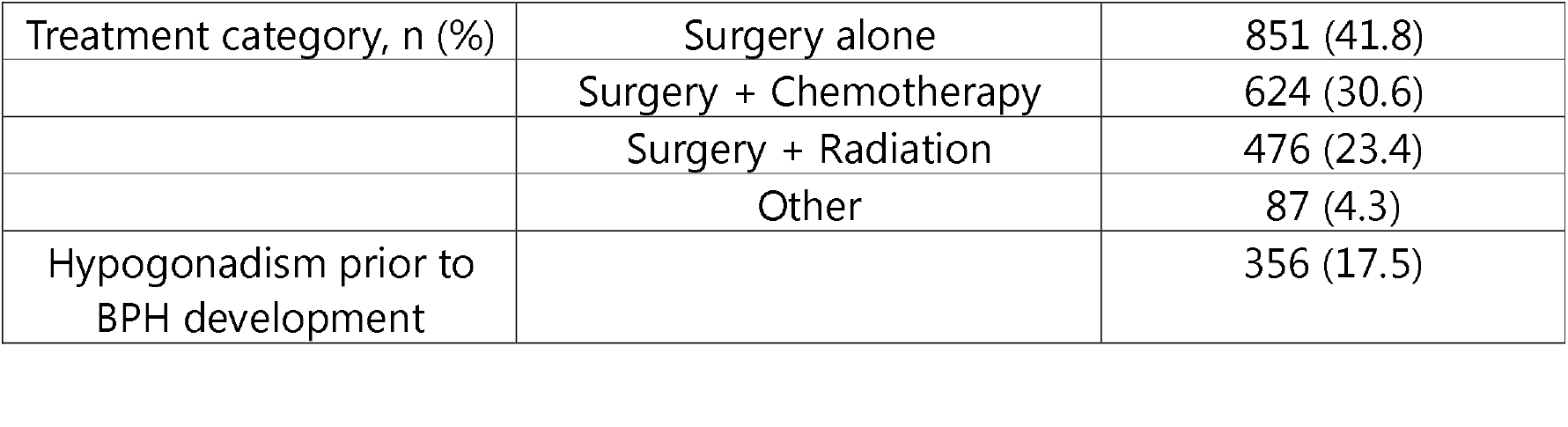
Demographics in Testicular Cancer Cohort.

**Table 2:** Cancer-specific factors and development of Benign Prostatic Hyperplasia.

| Variable |  | HR (95% CI) | p-value |
| --- | --- | --- | --- |
| **Univariable analysis** |  |  |  |
| Hypogonadism prior to BPH<br>(Ref: No hypogonadism) |  | 0.97 (0.74-1.26) | 0.9 |
| **Multivariable analysis** |  |  |  |
| Hypogonadism prior to BPH<br>(Ref: No hypogonadism ) |  | 1.05 (0.84-1.32) | 0.63 |
| Chemotherapy<br>(Ref: No chemotherapy) |  | 1.21 (0.94-1.55) | 0.13 |
| Age |  | 1.06 (1.05-1.07) | <0.001 |
| Race<br>(Ref: Asian/pacific islander) | African-American | 0.77 (0.26-2.24) | 0.63 |
|  | Native American | 0.81 (0.14-4.41) | 0.8 |
|  | White | 0.88 (0.32-2.37) | 0.8 |
|  | Other/<br>unknown | 0.67 (0.18-2.41) | 0.54 |
| Smoking status | Never | 1.02 (0.81-1.31) | 0.81 |
| (Ref: Current) |  |  |  |
|  | Prior history | 1.15 (0.89-1.49) | 0.26 |
|  | Unknown | 1.21 (0.91-1.59) | 0.19 |
| Decade of cancer diagnosis<br>(Ref: 1990-1999) | 2000-2009 | 1.78 (1.35-2.35) | <0.001 |
|  | 2010-2021 | 2.77 (2.02-3.79) | <0.001 |

Our study examined the incidence of BPH in TCa survivors, with a specific focus on the effects of chemotherapy and post-TCa hypogonadism. We found that, while BPH is a common condition in TCa surivors, neither chemotherapy nor hypogonadism were significantly associated with an increased hazard of BPH. Age at the time of TCa diagnosis was the primary factor influencing BPH development.

These findings align with existing literature, which identifies age as a well-established risk factor for BPH^2^. The absence of a significant association between chemotherapy and BPH is a novel finding, as it challenges the hypothesis that cytotoxic cancer therapies may contribute to prostate enlargement through inflammatory pathways^5^. Similarly, the lack of a significant association between hypogonadism and BPH suggests that testosterone deficiency alone may not be sufficient to drive the development of BPH in TCa survivors.

Our study isn’t without its limitations. As a retrospective analysis, it is subject to inherent biases, including potential misclassification of BPH and reliance on administrative codes for diagnosis. Additionally, the generalizability of our findings may be limited to veterans, who may differ from the general population in terms of health status and access to care.

Future research should explore the potential mechanisms underlying the observed age-related increase in BPH risk and investigate whether other factors, such as lifestyle or comorbidities, may interact with age to influence BPH development. More research is also needed to confirm our findings in more diverse populations and to explore other long-term health outcomes that TCa survivors face.

## Data Availability

Data from the VA is available through VINCI for VA investigators with appropriate institutional approval (San Diego VA IRB), and is not publicly available due to confidential patient records.

## Supplementary material

### BPH

- **ICD 10**: N40.1, **ICD 9**: 600.01
- **CPT codes:**
  - TURP: 52601
  - Laser enucleation : 52647, 52648, 52649
  - Lap simple prostatectomy: 55867
  - Rezum: 53854
- **Medications:**
  - Alpha blockers: terazosin (Hytrin), doxazosin (Cardura), tamsulosin (Flomax), alfuzosin (Uroxatral), and silodosin (Rapaflo)
  - Alpha-reductase inhibitors: Finasteride (Proscar) and dutasteride (Avodart)
  - PDE5 inhibitor: Tadalafil
  - Anticholinergic agents: Oxybutynin (Ditropan), Solifenacin (Vesicare), Darifenacin (Enablex), Trospium (Sanctura), Tolterodine (Detrol), Fesoterodine (Toviaz), Propantheline (Pro-Banthine)
  - Beta-3 agonists: Mirabegron (Myrbetriq) and Vibegron (Gemtesa)

### Hypogonadism

- **ICD 10**: E29.1, **ICD-9**: 257.2
- **Prescriptions**: exogenous testosterone (all formulations such as injectables, topical medications and subcutaneous implants)

